# Practices, norms, and aspirations regarding the construction, validation, and reuse of code sets in the analysis of real-world data

**DOI:** 10.1101/2021.10.14.21264917

**Authors:** Sigfried Gold, Harold Lehmann, Lisa Schilling, Wayne Lutters

## Abstract

**Objective:** Code sets play a central role in analytic work with clinical data warehouses, as components of phenotype, cohort, or analytic variable algorithms representing specific clinical phenomena. Code set quality has received critical attention and repositories for sharing and reusing code sets have been seen as a way to improve quality and reduce redundant effort. Nonetheless, concerns regarding code set quality persist. In order to better understand ongoing challenges in code set quality and reuse, and address them with software and infrastructure recommendations, we determined it was necessary to learn how code sets are constructed and validated in real-world settings.

**Methods:** Survey and field study using semi-structured interviews of a purposive sample of code set practitioners. Open coding and thematic analysis on interview transcripts, interview notes, and answers to open-ended survey questions.

**Results:** Thirty-six respondents completed the survey, of whom 15 participated in follow-up interviews. We found great variability in the methods, degree of formality, tools, expertise, and data used in code set construction and validation. We found universal agreement that crafting high-quality code sets is difficult, but very different ideas about how this can be achieved and validated. A primary divide exists between those who rely on empirical techniques using patient-level data and those who only rely on expertise and semantic data. We formulated a method- and process-based model able to account for observed variability in formality, thoroughness, resources, and techniques.

**Conclusion:** Our model provides a structure for organizing a set of recommendations to facilitate reuse based on metadata capture during the code set development process. It classifies validation methods by the data they depend on — semantic, empirical, and derived — as they are applied over a sequence of phases: (1) code collection; (2) code evaluation; (3) code set evaluation; (4) code set acceptance; and, optionally, (5) reporting of methods used and validation results. This schematization of real-world practices informs our analysis of and response to persistent challenges in code set development. Potential re-users of existing code sets can find little evidence to support trust in their quality and fitness for use, particularly when reusing a code set in a new study or database context. Rather than allowing code set sharing and reuse to remain separate activities, occurring before and after the main action of code set development, sharing and reuse must permeate every step of the process in order to produce reliable evidence of quality and fitness for use.

## 1. Introduction

As informaticists, epidemiologists, clinical researchers (not to mention regulators, health system administrators, software developers, and health economists), we believe patient data in clinical data warehouses (CDW) and distributed research networks (DRN), if properly analyzed, can yield transformational insights about patient populations and individual patients, and advance our knowledge of the biological, social, and institutional phenomena that affect patient health. Without delving into questions of data capture or the reliability of real-world data (RWD) or routinely collected data (RCD) analysis generally, this paper addresses the construction, validation, and reuse of a fundamental building block of RWD analyses [1].^1^

*The* fundamental building block of RWD analysis is, arguably, the electronic phenotype,^2^ the algorithmic expression of a clinical phenomenon of interest for the analytic task. The central focus of this paper is not the phenotype but the code set,^3^ which often serves as a phenotype’s primary building block. We recognize that coded data — data captured in the form of concept identifiers from controlled medical vocabularies — by themselves are seldom sufficient for accurately identifying clinical conditions in RWD [7,8]. Many phenotype algorithms incorporate uncoded data such as narrative notes and numeric observations and involve conditional and temporal logic. Nevertheless, vocabulary codes are generally the starting point for analysts designing phenotypes.

With RWD, data capture is not controlled by the researcher, and vocabulary codes are not always used as intended or expected. Each code represents a term or concept from a controlled medical vocabulary. The collection of terms in the code set are *functionally synonymous* — meaning that querying records that contain any one of these codes should ideally match all and only the patients who have experienced the phenomenon of interest [9]. In order to approach this ideal goal, the code set may need to be adapted to idiosyncrasies in the patient data. Therefore, purely *semantic* validation of a code set by experts in the terminologies used and the clinical subject matter may not be considered adequate; *empirical validation* of patient data may also be needed.

Discussions of code sets and their validation and reuse in RWD applications have, we suggest, been muddled in prior literature because they do not distinguish the RWD context from applications where some control over data capture is possible, such as common data element (CDE) design and clinical quality measures (CQM), in which purely semantic validation may be sufficient. Empirical synonymy is not the same as semantic synonymy, so the validity of a code set in a phenotyping application depends on the data being analyzed, the patient population, local coding practices, clinical workflows, and more.

Concerns regarding the quality of code sets and the challenges faced by code set developers in identifying and selecting the set of codes best fitted to their clinical intention and analysis goals have inspired the development of code set authoring and sharing platforms [10–12] and other scholarly and practical efforts to champion and facilitate code set reuse [13–25].

Despite the development of code set repositories and other tools, concerns around code set quality persist [19,26–28,9]. These concerns are addressed most fully in a 2017 paper by Williams, et al., “Clinical code set engineering for reusing EHR data for research: A review” [26]. It is the first paper to address code set engineering and reuse generally and to call out code set use in RWD research as a worthy topic of sustained study. It provides nomenclature, a consolidated articulation of published knowledge on code sets, and a valuable catalog of recommendations for advancing technology for managing code sets. It serves as a touchstone throughout the current paper, a starting point for elaboration and critique. We will abbreviate its name below as CCSE.

Though CCSE focuses specifically on phenotyping applications, it does not call out the unique challenges these applications present in the areas of validation and reuse. In order to understand the real-world practices surrounding the use of code sets, we designed and conducted a field study of epidemiologists, clinical researchers, or informaticists and others who work with RWD. We conducted an online survey, semi-structured interviews with a subset of survey participants, and observation where possible of participants performing actual work to create code sets.

Our study design intended to characterize the practice of code set development: the variety of institutional settings and analytic goals of code set developers, the data and software available to them, how contextual factors affected code set development and use. We were attuned to whether the concerns and processes shaping code set development were as heterogeneous as the contexts or not.

While there appears to be universal agreement that crafting high-quality code sets is difficult, our participants held very different ideas about how this can be achieved and validated. A primary divide exists between those who rely on empirical techniques using patient-level data and those who only rely on expertise and semantic data. Code set development practices relying on patient data must grapple with problems around private health information (PHI), software configuration, and questions of cross-database validity. Rigorous validation may require PHI, but the privacy and specificity of patient databases constrain possibilities for reuse. These tensions, we propose, have stymied past efforts to promote meaningful, widespread reuse of code sets for phenotyping.

CCSE breaks code set management down into four phases: construction, validation, sharing, and reuse. These terms are intuitively descriptive, and we continue to rely on them, but the practices found in our study could not easily be classified into one of these phases and necessitated the formulation of a structured process model, which is able to account for observed variability in formality, thoroughness, resources, and techniques.

Between the requirements planning and naming of an initially empty code set and the decision that it is complete, the diverse array of tasks involved in assembling and refining it comprise, according to our model, an iterative cycle of (1) code collection, (2) code evaluation, and (3) code set evaluation. When this cycle ends, the code set is (4) accepted and ready for use. These steps are sometimes followed by (5) reporting — for publication, documentation, or stakeholder requirements — on the methods and results of creating and validating the code set.

We analyze the real-world practices and ideas of code set developers and synthesize these with accounts of code set validation and reuse in the literature, as well as our own professional experience. Our findings and recommendations problematize understandings of code set validation and reuse while offering novel perspectives and approaches to capturing and sharing evidence to support trust in the quality and fitness for use of code sets and metadata being considered for reuse.

## 2. Methods

The insights presented in this paper were derived from a triangulation among three primary data sources: a survey of medical informaticists, contextual interviews with leading code set practitioners, and observations from decades of community participation by the first three authors. The initial 21-question, web-based survey (see Appendix A. Survey questions) investigated participants’ experiences using clinical code sets in the analysis of RWD.

Recruitment focused on professionals with experience using code sets in the analysis of RWD. These participants were identified through the first author’s professional networks. Given the variety and inconsistency in nomenclature for RWD analysis elements and processes, questions were carefully balanced to capture differences in interpretation and use.

Of 36 completed surveys, 15 survey respondents were invited for a follow-on semi-structured interview, conducted as a videoconference by the first author. The purpose of the interviews was to explore their code set authoring and reuse practices. Within this set four were able to participate in extended contextual interviews where they were able to demonstrate their tools and processes for developing code sets via screen share.

We performed open coding and thematic analysis on interview transcripts, interview notes, and answers to open-ended survey questions.

The first author has 13 years of experience as a medical informaticist and research programmer working with RWD and clinical code sets. She has contributed to numerous projects and key communities in this space, including the Army Pharmacovigilance Center, Observational Health Data Science & Informatics (OHDSI), the National COVID Cohort Collaborative (N3C), and the American Medical Informatics Association. These experiences provide an orientation to the practices explored in this study and a sensitivity to our participants’ goals and concerns.

The qualitative data collected from the surveys, interviews, and observations was content coded in NVivo through a process of analytic induction informed by the first author’s personal experience in the field. Codes and emerging themes were iteratively developed with co-authors. This human subjects research was reviewed and approved by the University of Maryland IRB (#**1405794-8**).

## 3. Results

### 3.1. Participant demographics

Seventy survey invitations were sent out. Of the 49 responses, 36 were completed enough for analysis. Our response rate was 64% and completion rate was 47%.

Table 2 shows the diversity of our sample population in terms of relevant demographic and work environment characteristics. Participants held an array of degrees. The listed disciplines reflect professional focus, not necessarily training.

**Table 1.**
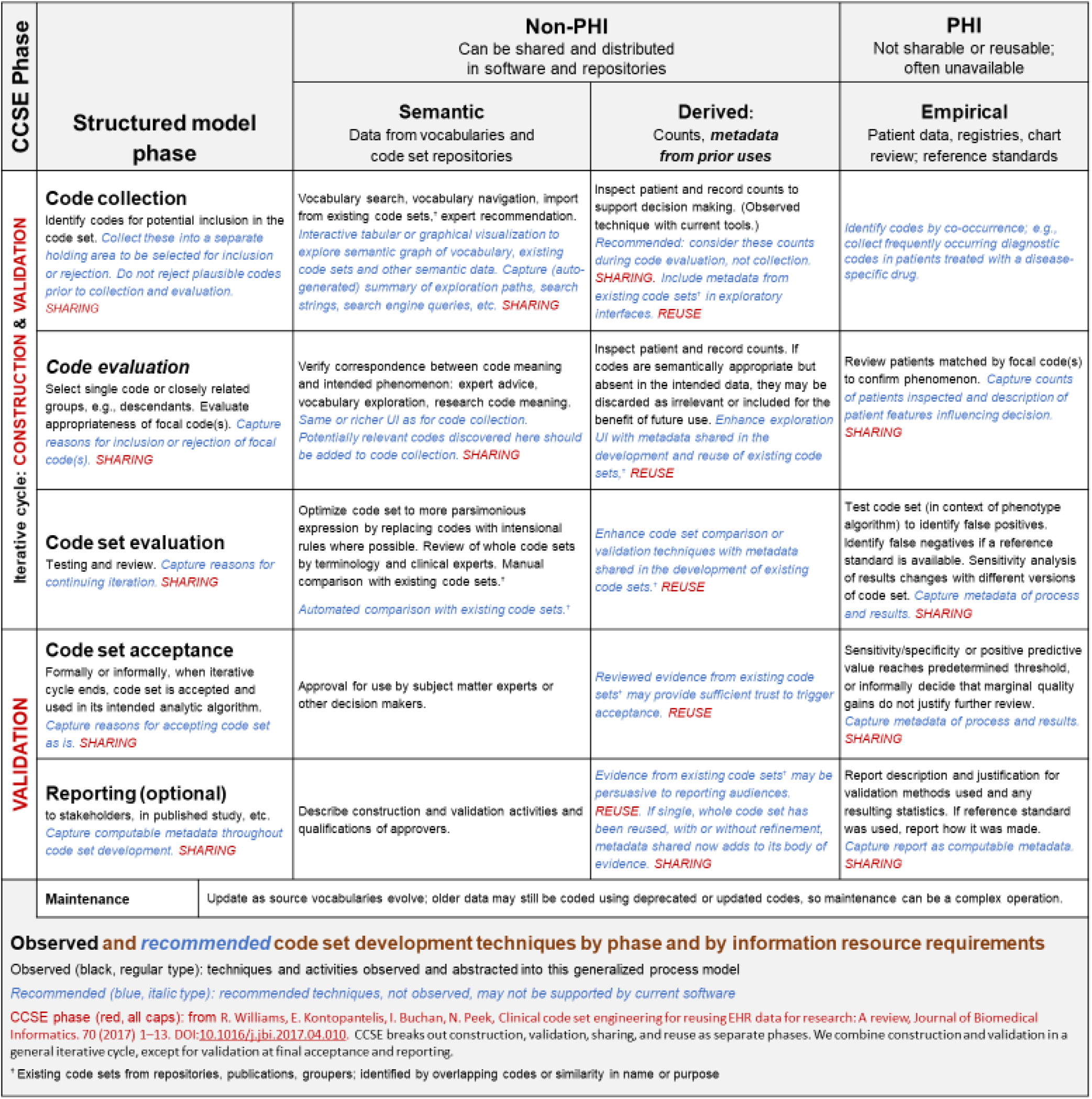
Structured process model and taxonomy of code set development, with recommendations

**Table 2.**
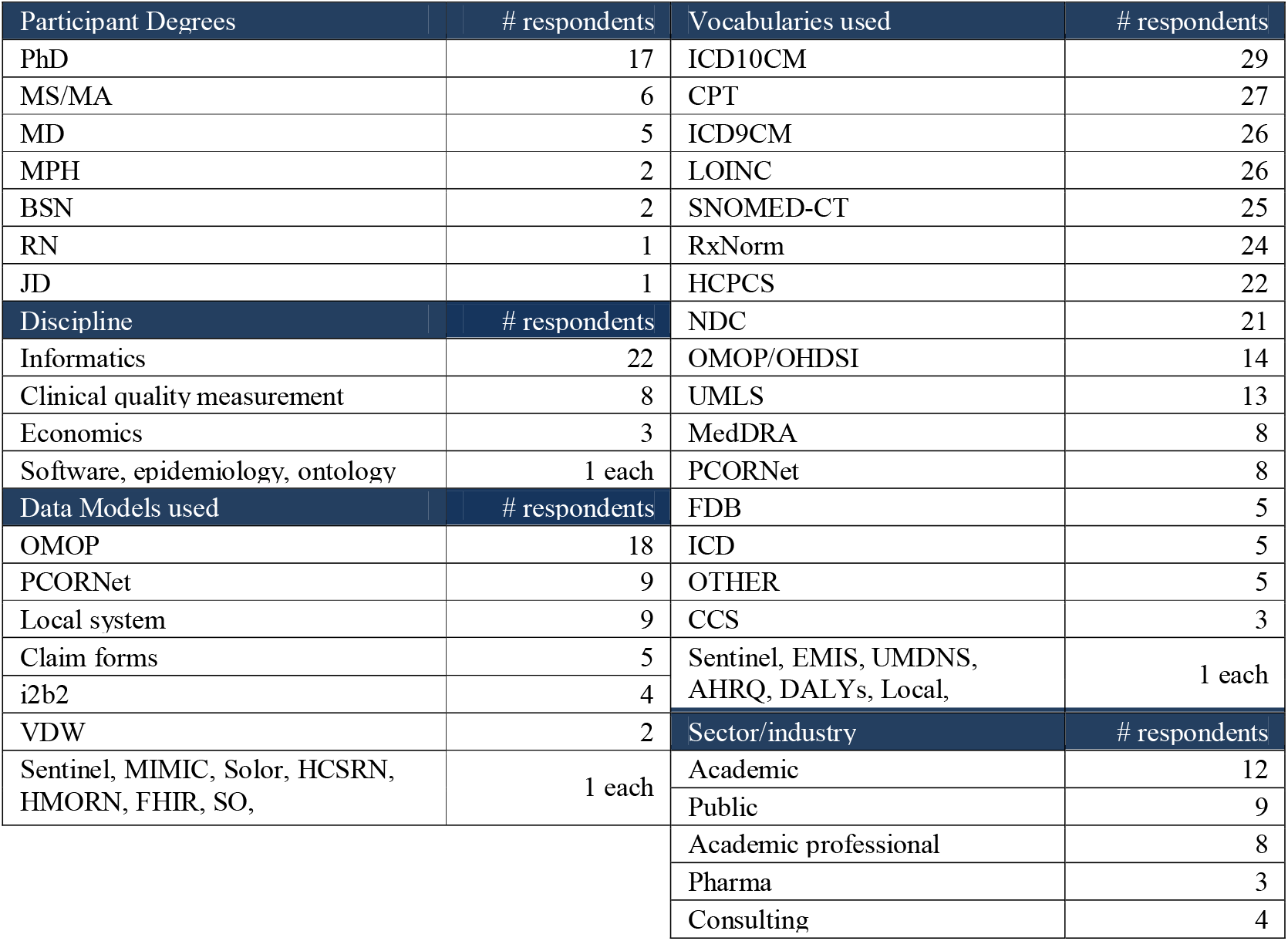
Participant demographics and work environments

### 3.2. Variability

We found significant variability in code set development practices and norms along numerous dimensions, listed in Table 3.

**Table 3.** Dimensions of variability

|  |
| --- |
| The formality and rigor of code set development workflow and validation |
| The types and degree of expertise of those managing code set development and the larger analytic process |
| the degree of integration between these processes and their resulting components, such as study protocols, phenotype, cohort, and analytic variable definitions, etc. |
| The information and resources used in code development, such as terminology resources, code set repositories, CDWs and software tools and platforms |
| The effort spent in exploratory analysis of patient data and terminology resources |
| The formality and types of reports generated, if any, of assessment processes and measures. |

The following quotes encompass the range in the formality of validation we saw in the answers to SQ16 (“How do you verify that you have selected the best codes for representing a clinical concept in your analyses?”):^4^

> First conduct discussion with clinical experts; Second, evaluate coverage of clinical concept in a data set; Third, perform random chart review to help detect if presence of code indicates disease (*P16*)
>
> Depends on the purpose and whether we are aiming for sensitivity or specificity. It may be chart review, or comparison with other value sets. (*P34*)
>
> We are usually given the value set (P12)
>
> By my background knowledge (P23)

#### Contribution 1.

*Strategies for gaining confidence in code set accuracy vary widely in their apparent formality and rigor, from systematic clinician chart review to analyst spot checking of patient records, from Delphi techniques to vaguely defined sanity checks based on vocabulary review or expert knowledge. (See* *Sections 3.8.1* *and* *3.8.2*.)

The variety we see in our participants’ answers is such that we count all these strategies and techniques as “validation”. Validation as described in CCSE [26] may encompass a reporting component which may include a description of validation methods employed or measurement statistics such as sensitivity, specificity, and/or positive predictive value (PPV).

### 3.3. Validation for code set reuse

There is a wider literature on RWD or RCD reporting guidelines [8,29–34] with specific recommendations on what aspects of study design, cohort selection or phenotyping, and code selection and the validation of these should be reported. The focus on how study design and validation should be reported generally stops short of saying what validation methods should be used. This literature is concerned with reporting individual studies in the literature and does not seem to address questions of how study elements like phenotype definitions and code sets and their validation could be shared to meet the needs of potential re-users.

Technical requirements for reuse are covered in “The FAIR Guiding Principles for scientific data management and stewardship” [35], FAIR being the acronym for Findable, Accessible, Interoperable, Reusable. P07 — a professor of medical informatics specializing in semantic interoperability, terminologies, information modeling, and standardization — offered as a best practice to facilitate discovery and reuse of code sets, “Representing them as FAIR objects” (SQ18). And as a change they would like to see in the design of code set management tools, “Enable use of and contribution to repositories of FAIR objects” (SQ20).

Beyond the requirements for making reuse of code sets technically possible — already accomplished by the Value Set Authority Center (VSAC) [10,11] and other tools — meaningful reuse will also depend on potential code set re-users being able trust its quality and fitness for their use, which will require access to evidence regarding its validation and purpose. Alper, et al., 2021, “Categorizing metadata to help mobilize computable biomedical knowledge,” expands FAIR to FAIR+T to encompass trust [25]. Though that paper offers example metadata for value sets and other computable objects, it does not address the difficult problem of sharing validation evidence (see Sections 3.12 and 4).

### 3.4. Reference standards and formal validation practices

We found considerable correspondence between the validation methods described in CCSE and those described by our survey and interview participants. Among the “several processes encompassed by code set validation,” CCSE particularly highlights two: (1) “internal validation typically with a clinician…when the final code set is examined to confirm that all the codes included are relevant to the concept of interest. This is an important step to reduce type I errors, where an incorrect code is wrongly included,” and (2) “creating a list of synonyms, using a code hierarchy and searching iteratively…, an important part of the code set construction process, [for] reducing type II errors, where a valid code is incorrectly omitted [26, p. 8].

Type I errors, i.e., inappropriate codes are included, tend to increase false positives; and type II errors, i.e., appropriate codes are omitted, tend to increase false negatives. One participant mentioned that when codes are added to a code set to correct type II errors, the intent is to transform false negatives into true positives, but the new codes may also transform true negatives into false positives.

The only validation method capable of yielding a full 2×2 table (see Table 4) and producing sensitivity and specificity statistics is by comparing matched records against a reference standard – a sample of records already tagged as presenting the phenotype or not. This approach is offered as a recommended guideline in the RECORD Statement [31, p. 4, items 6 and 7] and in Callahan, et al. 2020 [34, p. 10].

**Table 4.** 2×2 with MRA-generated reference standard

|  | Condition present in database | Condition absent in database |
| --- | --- | --- |
| Algorithm positive | True positives (TP) | False positives (FP) |
| Algorithm negative | False negatives (FN) | True negatives (TN) |

After recommending vocabulary navigation and sensitivity analysis, CCSE [26, p. 8] describes using “a gold standard measure such as manual notes review” as a “more complete way to validate a code set” but says that fully deidentifying patient data is difficult and costly and, without deidentified data, code set developers may not be able to use patient data at all.

### 3.5. Obstacles to formal validation

Every one of our 36 survey respondents reported that their studies use patient data. Nevertheless, only a small subset had access to deidentified data. Of the 32 who answered the open-ended validation question (SQ16), only nine indicated using patient data for validation. In one interview, P31 said it would have been cost prohibitive to do chart review even if they had had the data. In a follow-up interview, they explained that systematic chart review is their usual practice in RWD studies, but this study compared code set results with patient-reported outcomes, so their usual practice was not absolutely necessary.

Reference standards are generally created by medical record abstraction (MRA), a systematic process of chart review conducted by qualified clinicians or trained abstractors. Many of our participants mentioned chart review, sometimes implying such a formal process, other times implying no more than cursory sanity checks on a handful of records; usually by a clinician, though sometimes by an analyst without clinical training, such as an informaticist or epidemiologist. Even when conducted according to recommended guidelines, MRA is a complex, multifaceted task, prone to subjectivity, with “high and highly variable discrepancy rates.”[36]

Most participants were familiar with validation by MRA-generated reference standard, and primarily discussed its impracticality and other failings. The problems with this method were more than data unavailability, cost, and doubts around the accuracy of MRA. It extended beyond questions of reference standards to address whether code sets have independent meaning at all:

> I don’t think you can separate notion of concept set from cohort, because concept sets are devoid of data, but its application of concept sets, alongside temporal logic, that becomes instance to find persons in dataset. (P05-SQ18)

If P05 is correct, the only way to “validate” a concept set would be by validating its containing cohort or phenotype algorithm. This view was raised by only two participants, but we find it persuasive, and, when probed, other participants tended to agree with it.

#### Contribution 2.

*A code set cannot be empirically validated except in the context of its containing algorithm*.

At the same time, however, our participants and others [9,12,26,27] seemed to hold an unspoken assumption that valid and validated code sets exist, and that code sets should be validated and made available for reuse. These assumptions may be based on a conflation of code sets in RWD analysis and value sets in other contexts.

> I work closely with a clinician who can help me establish the initial list of synonyms and validate the final code list. (P43-SQ16)
>
> We use validated codesets when possible(P04-SQ16)
>
> [Reusable code sets] would be helpful [so that] I don’t have to do this on my own every time…[B]ecause it has been created by a collaborative team that’s known for creating value sets, I would know that, “Oh, this has been extracted or they got it from a paper that has been vetted and validated and you know it’s a legit paper.” I would use that. (P09-interview)

### 3.6. Permissible value sets and analytic code sets

“Clinical code set”, “code set”, “concept set”, and “value set” are generally used synonymously (see [26, pp. 2, 9] and footnote on p. 1). Different participants in our study preferred one term or another but appeared to use them interchangeably. The focus of our study on the use of code sets in the analysis of real-world data was made clear to our participants in the survey text and during interviews. Nevertheless, many made no distinction between these and permissible value sets, i.e., lists of values (codes) constraining the permissible contents of a data element. We have been unable to find the distinction between permissible value sets and analytic code sets articulated in the literature, though their conflation seems to cause confusion, especially around validation.

#### Contribution 3.

*A permissible value set constrains a data element or domain to the universe of semantically distinct terms it can contain. In contrast, an analytic code set is meant to hold a set of functionally equivalent terms, any of which indicates an instance of a more general concept*.

Value sets for CQMs are a special case where the terms are functionally equivalent, but the CQM authority dictates which terms will be accepted. Authors may sometimes consult EHR data in quality measure design, but these measures tend to be applied regionally or nationally, not to any specific database, and it is the responsibility of the institution running the CQM to assure that data are coded to match the CQM’s value sets. For that reason, we classify these as permissible value sets and CQMs as non-RWD applications.

As a contrived illustration, a permissible value set for gender might be: Female, Male, Other, Refused to answer, Unknown. Each term is distinct. An analytic code set for female might be (assuming case-insensitivity): female, f, fem; all synonymous and interchangeable for query purposes. And an analytic code set for non-male might be: female, f, fem, other, non-binary. Clearly, f, other, and non-binary are not synonyms, but in reference to the concept of non-male, they are equivalent.

There are many ways to validate value sets in contexts outside RWD analysis, some of which we explored in the study but are out of scope for this paper. According to a 2012 paper, “Quality evaluation of value sets from cancer study common data elements using the UMLS semantic groups,”

> With the importance of value sets gradually being recognized by the clinical research community, standardization of value sets is becoming imperative, as it can enable comparison across disparate datasets and facilitate reuse of well-defined value sets to advance clinical research studies [5].

The importance of facilitating and eventually standardizing code set reuse, recognized in this non-RWD context, is also recognized by some in the RWD or phenotyping community. It is espoused explicitly in CCSE [26] and Springate, et al. [12]. It appeared to be taken for granted by many of our participants. Prior to conducting this study, we had recognized and described some of the obstacles to reuse [9,27] but had not yet grasped that reuse depends on communicable validation reporting.

Even setting aside the issue of costly reference standards, how is it that so many continue to believe in the value of code set reuse in RWD if code sets on their own cannot be validated? Is it because they *can* be validated in other contexts, so people just do not realize that RWD is somehow different? Is it because code sets are effectively phenotypes unto themselves and can be validated on their own? Or that when people endorse the value of code set reuse, they are really talking about phenotype reuse?

### 3.7. Prescriptive and descriptive code sets and validation

There is a difference between RWD and non-RWD contexts that corresponds, we posit, to a difference in perspective about what code sets are:

- A code set is an expression of clinical meaning based on the clinical understanding and classifications built into medical vocabularies, it comprises the set of codes that *should be used* to indicate the clinical phenomenon of interest *in any database* of electronic health records.
- A code set is a collection of codes that *have been used, correctly or not*, to indicate a clinical phenomenon of interest *in a specific database*.

In lexicographical or grammatical terms, the first perspective is *prescriptive*, and the second is *descriptive*. While terminological prescriptivism in natural language is considered unscientific and pedantic [37], it is, of course, the foundational purpose of standardized medical vocabularies and arguably the foundational practice of medical informatics generally [38–41].

Exemplifying a prescriptivist orientation, Winnenburg, et al. 2013 defines “quality criteria for value sets in clinical quality measures”:

> A value set should contain all the relevant codes for a particular data element. […T]he value set is expected to be rooted by one concept and to contain all the descendants of this root concept. [14, p. 1]

By this view, any need for a code set containing more than a single code and its descendants would signal a failure in vocabulary design or in code assignment. This view could not be farther from the descriptivist view shown more commonly in the phenotyping community, as exemplified in the following:

> I do some analysis of result set produced by queries with a given value set, using descriptive statistics and visualization. Sometimes I will also look at data missed by the value set to see if there are additional revisions that I should make. (*P17-SQ16*)

The binary classifications we have outlined, between non-RWD and RWD validation contexts and prescriptive and descriptive terminology use, imperfectly line up with two more binary classifications: between semantic and empirical validation techniques and between public and private data.

### 3.8. Semantic techniques are based on sharable data, empirical on protected

In the terms of some of our participants, the focus on meaning is called *semantic* and the focus on use is called *empirical*.

> Lexical search,^5^ semantic exploration (navigate OHDSI vocab), empirical assessment thru characterization, and clinical expert review. (P05-SQ16)

We give the terms semantic and empirical a precise, practical definition, which may seem idiosyncratic, but preserves the denotative meaning understood by our participants, clarifies some edge cases, and concretizes the distinction to provide further practical benefits.

#### Contribution 4.

*“Empirical” techniques are those requiring reference to detailed patient-level data. “Semantic” techniques are those requiring anything else, including standard vocabularies, vocabulary mappings, ontologies, existing code sets, and groupers*.

Patient-level data can seldom be fully deidentified and generally qualify as protected health information (PHI). There can be crucial differences in the data elements available and how these are structured and accessed across institutional and database contexts. While code sets target coded data, empirical validation, particularly with an MRA-generated reference standard, is likely to require clinical notes, lab results, images, and other non-coded data. Even if these data will be available when the full study is run, they may not be available to code set developers. Some data constraints, limitations, or complexities may not be discovered until deep into a project. All told, the use of empirical techniques is often encumbered with high resource demands and legal, institutional, and technical hurdles [42].

#### Contribution 5.

*Empirical, patient-level data are generally private, protected, institutionally idiosyncratic, missing data elements that may be needed, expensive and cumbersome to access and use*.

#### Contribution 6.

*The purpose of empirical techniques in code set development is to assure that the code set accurately retrieves patient records indicative of the clinical phenomenon in question when applied to the database(s) analyzed. (See* *Section 3.8.2*.)

#### Contribution 7.

*Empirical data and techniques may be indispensable for validation depending on analytic and reporting requirements*.

Semantic data — in the form of digitally encoded vocabularies, groupers, code set repositories, etc. — though they may be encumbered by licensing constraints and complexities (e.g., around version synchronization), they are not burdened with the same kind of privacy issues that are always present with patient data. Though medical terminologies may differ greatly in structure and classificatory principles, dealing with these differences is mitigated for users of curated and harmonized vocabulary systems such as UMLS [43], the OHDSI/OMOP vocabulary systems [2], and CDMH [44,45].

The small portion of participants discussing empirical validation may be evidence of the difficulty of performing it. It is also striking that more technically proficient participants who discussed writing analytic code (in SQL, Python, etc.) all had some degree of access to empirical data, but — in the service of code set development — only used semantic data.

#### Contribution 8.

*Semantic data, in contrast to empirical data, can be distributed and used without exposure of PHI*.

We call attention to the association of semantic with sharable data and empirical with protected data believing that it sheds light on challenges involved in designing code set management tools and techniques relying on empirical data, as well as on opportunities to make more effective use of semantic data in tool design.

#### 3.8.1. Semantic techniques and data

##### 3.8.1.1. By imputed authority

The validation techniques most frequently mentioned by our participants are of two types: (1) consultation with or review by terminology and domain experts and (2) use of or comparison to existing code sets found in published literature, code set or phenotype repositories, and published code groupers. We could describe both types as “validation by authority”, as long as we recognize that the authority is reputational, it can only lend credibility to a code set specification. These experts and respected sources are trusted to know how terms are used in data, but they have no power to enforce it.

Following are several more answers to SQ16 (see Appendix A. Survey questions) that emphasize validation by authority. In some cases, the authority is a person:

> Oversight with terminologists, clinicians (P19)
>
> We have a clinical expert review the concept sets. (P26)
>
> We discuss with our coding panel, a group of experts that give us advice and feedback. (P21)
>
> Depends on domain. Usually verify with clinical SME when available, or conventions within a network (P44)

In other cases the authority takes the form of existing code sets:

> Examine the literature for validation studies. (P41)
>
> previous published results (P01)
>
> Prior literature, technical reports (P36)
>
> Verifying against the current billing practices (claims) or validated phenotypes (P06)

Validation by authority can occur along a continuum of formality and rigor. At the more formal end, we had one participant (P31) describe systematic code set development, in a Delphi-like process with inter-rater review. They weren’t satisfied with the results and were able to find and fix problems with follow up efforts. They “shopped their code sets around” to specialists “to see if there were relationships that we could use to increase sensitivity and specificity.” They also had an analyst find problems by coming up with patient record scenarios that would produce false negatives and false positives. After all this, they believed they would have been better off with chart review or sensitivity analysis, but the project lacked sufficient funding.

Expert knowledge and existing code sets can both serve as sources of authority; regardless of how authoritative they are considered, they serve to bolster confidence in the appropriateness of the codes they suggest or contain.

#### 3.8.1.2. By computable semantic knowledge

Several participants discussed their use of vocabulary browsers in the process of constructing code sets. Some used vocabulary browsers provided by the vocabulary maintainer, others used third-party browsers such as the OHDSI ATLAS concept set builder [46] or OHDSI’s Athena [47] tool for browsing and downloading vocabularies, one used the vocabulary tree built into i2b2 [48]. All these tools allow hierarchical navigation of vocabulary structures.

We observed P14, an epidemiologist in the drug safety, real-world evidence department of a large pharmaceutical company, using ATLAS, the web interface to the OMOP common data model and OHDSI tech stack. P14 narrated their ATLAS vocabulary search for generic or brand names for a particular drug:

> Precise ingredient, that’s not what I want. There it is. Oh, I just saw it go by…So then I would put that into my concept set, and I would start taking a look at what’s underneath of it, like in RxNorm at least.

And later, on a related task:

> I can see the brands, but I actually can’t really use ATLAS to split them up very easily. So, to be honest, I would probably need to go out and write some SQL at this point.

One participant used GetSet [24] to iteratively refine a code set using Read Codes. With GetSet, an initial string search immediately populates the code set with terms containing the search string and with all the descendants and synonyms of those codes. If the resulting list contains terms inappropriate to the code set, search strings that match them can be added to an exclude list.

Another participant wrote Python scripts to download and compare a number of depression code sets from VSAC to find the most commonly used codes and to consider whether less commonly used codes should be included in a code set meant to cover depression generally.

> There are several value sets that define a diagnosis of depression, and so my first questions are, okay, why are there multiple value sets that do this? What are the differences? And then, as we get further into the process with the clinical experts on using, reusing, or expanding a value set I want to understand what that overlap is…
>
> [I]n the ideal world we’d resolve to an existing value set that we could reuse but there’s many value sets for similar concepts for a reason, and so it may be that we do need to create a new value set based off of an existing one… I’m sitting here wishing that there was an explicit acknowledgement by one of these authors that, oh yes, I know that this bipolar disorder grouping value set existed. I was aware of it. We reviewed it, and we created this one because dot, dot, dot. (P16)

Though many participants mentioned starting code set construction with string search in vocabularies, none considered it sufficient by itself for producing a code set. One participant suggested, however, that many researchers do think it sufficient:

> Sadly, most investigators still use string pattern matches for their work and don’t do ANY analysis of the coded values…I usually have to do cursory checks of the resulting codes and quite often find obvious issues…manual review with knowledge of the coding system and domain is critical. I usually find a trusted expert (ex: pharmacist or ontologist) and then review the code sets manually based on the intent… (P24-SQ16)

### 3.8.2. Empirical techniques and data

Though MRA-generated reference standard may be the canonical empirical validation technique, our participants discussed a range of empirical techniques for testing their code sets: doing spot check inspection of patient records, slightly more systematic chart reviews or MRA, and use of record counts, either for individual codes (with or without descendants) or for whole code sets. The following quotes come from faculty and staff at academic medical research centers with their own CDWs.

> First conduct discussion with clinical experts; Second, evaluate coverage of clinical concept in a data set; Third, perform random chart review to help detect if presence of code indicates disease (*P16-SQ16*)
>
> Chart Review. Some internal checking of codes against expected lab results, vital measurements, patient histories, etc. Ex. Diabetes codes should associate with histories of certain blood glucose measurements or A1c. (*P28-SQ16*)
>
> We benchmark across datasets in the federated analysis (P10-SQ16)
>
> Test in real world data (P41-SQ16)

More common than MRA-generated reference standard is another form of systematic chart review: performing chart review or MRA on a random sample of the records matched by a code set or phenotype to identify false positives. This procedure allows reporting of PPV, but not sensitivity or specificity. A common attitude, reflected in CCSE [26], is to rely on semantic techniques like vocabulary navigation or culling through existing code sets, for coverage, for avoiding false negatives; and to rely on empirical techniques like random chart review for correcting false positives and increasing PPV.

One participant (P12) described a form of sensitivity analysis in which the entire study was run using a full range of plausible code sets. If results remained stable across these, any differences in cohort composition would be considered immaterial.

PheValuator [28], mentioned by two participants, algorithmically produces a “probabilistic gold standard” to be used in validating phenotype algorithms and code sets. It is given two starting code sets, one perfectly sensitive, the other perfectly specific (as well as these can be determined.) Patients matched by the specific code set are considered positive cases, patients *not* matching the sensitive code set are considered negative cases. The PheValuator algorithm is trained on samples of these and assigns probabilities to remaining patients rather than tagging them as positive or negative.

### 3.9. Refining versus validating code sets

Without a reference standard the only digital evidence of the clinical phenomenon or phenotype being present or absent is what can be found in the CDW, either by ad hoc forays into the data or, more likely, by running the code set through database queries or the algorithm the code set is being designed for. If a reference standard is available, the set of records tagged in it can be run through the phenotype algorithm and a 2×2 table (Table 4) can be constructed. With all four cells of the table at hand, sensitivity, specificity, and PPV can be calculated and reported.

Without a reference standard to identify false positives and false negatives, however, misclassifications must be discovered by various semantic or empirical approaches to identifying them. With sufficiently sized random chart review, researchers may feel confident they have identified false positives. With true and false positives at hand, PPV can be calculated and reported. This approach will not help in identifying false negatives, as illustrated in Table 5.

**Table 5.** 2×2 with random chart review on positives

|  | Condition present in database | Condition absent in database |
| --- | --- | --- |
| Algorithm positive | **TP** | **FP** |
| Algorithm negative | **FN = <unknown>**<br>Only available as discovered | **TN = 100% - FN** |

If neither reference standard nor random chart review are used, none of the 2×2 table cells will be known (Table 6). Even supposing there is enough evidence somewhere in the database to support a determination of whether any patient has or does not have the condition, the evidence available to the code set developer consists of (1) what the algorithm (or code set) matches, and (2) whatever else they can determine or surmise by the variety of techniques they are able to deploy. As false positives or negatives are discovered, the developers will presumably modify the code set or phenotype. If code set errors are discovered by semantic means or otherwise without allowing for systematic empirical confirmation, its authors may assume, with or without justification, that correcting these errors will improve its accuracy, but they will not be able to quantify that improvement.

**Table 6.** 2×2 with other methods

|  | Condition present in database | Condition absent in database |
| --- | --- | --- |
| Algorithm positive | **TP = 100% - FP** | **FP = <unknown>**<br>Only available as discovered |
| Algorithm negative | **FN = <unknown>**<br>Only available as discovered | **TN = 100% - FN** |

#### Contribution 9.

*Editing a code set consists of adding and removing codes. Codes are added in order to correct type II errors, eliminate false negatives, and increase sensitivity, but may inadvertently introduce false positives. Codes are removed in order to correct type I errors, eliminate false positives, and increase specificity, but may inadvertently introduce false negatives*.

#### Contribution 10.

*When there is no formal acceptance procedure (e.g*., *reference standard or clinical and terminology review by subject matter experts), “validation” efforts are indistinguishable from code set quality refinement efforts. Whether or not a highly refined code set (or phenotype) produces more accurate results than a formally validated code set, its accuracy will not be measurable. Some experts have more confidence in rigorous refinement efforts than in the statistically reportable results of MRA-generated reference standards*.

When we asked participants about the value of documenting refinement efforts, they expressed enthusiasm, but affordances for accomplishing this can be found in few if any available tools. (See Section 4.)

### 3.10. Sharable derived data: an “empirical semantics”

In addition to validation methods based on patient-level data, several participants discussed term usage counts as important to their code set development processes.

> Compare them to published literature [to] determine the presence/absence of codes in our target datasets (P33-SQ16)

Through ATLAS, P14, the pharmacoepidemiologist described above (p. 14), could navigate all the vocabularies relevant to their analysis and could view patient profiles and run characterization reports for millions of patients. The only use of the empirical data they mentioned, however, was to eyeball the patient record counts of concepts they considered including in their concept sets. Describing researchers involved in PEDSNet [49] they said

> Patient-reported data is like their holy grail of trying to understand what’s going on with the patients…But the vocabulary helps, and ATLAS helps a lot because…it shows if you use this concept set, you are going to find a decent amount of people that get associated to this diagnosis.
>
> I usually sort [codes] by how much evidence they have, and I can see that there’s a lot of evidence associated to this kind of top-line SNOMED code.

Given all the shortcomings of RWD [c.f. 50, p. 1], few assumptions can be made in their analysis, and it is always wise to check one’s code sets and algorithms against empirical data, or even, as P14 says about PEDSNet researchers, supplement them with more reliable data in the form of patient-reported outcomes. Empirical data and techniques are essential to the validation of code sets and algorithms, though the only empirical data we observed participants using were patient and record counts. Unlike patient-level empirical data, counts can be shared publicly.

#### Contribution 11.

*A third category of shareable data derived from PHI includes term usage counts and other aggregate data as well as evidence and metadata culled from previous analyses*.

We label these data “empirical semantics” with the idea that they will form a supplement, possibly almost an alternative, to vocabularies and authority-based semantic data; a semantics not of intended meaning but of found meaning. PHI cannot be shared, but useful derived information can be. For instance, a study team’s code set and phenotype developers could be aided considerably by learning that diagnostic code A in database B only indicated an actual diagnosis of A in 60% of B’s patients, while indicating suspected A (and possible cause for lab testing) in 37% of patients, and erroneous data in 3%.

Patient counts are the primary (if not the only) currently available form of data we classify as empirical semantics. Section 4.1 suggests other methods for capturing valuable data through the code set development process itself. Some of these data can be derived from PHI without exposing PHI.

### 3.11. A structured process model

CCSE describes four phases of code set “management”: construction, validation, sharing, and reuse [26, pp. 7-9]. These make sense intuitively: the author constructs the code set, validates it, shares it, and then others come to reuse it. We see from our evidence as well as remarks in CCSE itself that there is perhaps no part of the construction phase that would not also be considered part of the validation phase. Repeating P05-SQ16, their process for verifying the accuracy of their code sets is: “Lexical search, semantic exploration…, empirical assessment…, and clinical expert review.” CCSE also includes all aspects of construction in the validation section as beneficial for reducing type II errors.

Our process model reflects our observations as well as our analysis, synthesis, and recommendations — the recommendations are shown in italic, blue type in Table 1**Error! Reference source not found**.; the remainder of that table presents a useful abstraction of the various activities that can occur in developing an analytic code set. Though our model does not pretend to contain an exhaustive account of all techniques used in code set development, we have aspired to offer a taxonomy capable of classifying any technique^6^ into one of five sequential phases (three of which form an iterative cycle) and three categories of information source.

Insofar as our model reflects our findings on real-world code set development, it does not address sharing or reuse as our participants provided few examples of those activities. Code set development consists primarily of an iterative construction/validation cycle of code collection, code evaluation, and code set evaluation. We are not claiming that our participants demonstrated or told us about the kind of structured process steps our model describes. A code set developer is not likely to think, “First, I will collect codes, then I will evaluate them for inclusion individually or in groups, then I will evaluate the whole code set, after which I shall iterate on those three steps until satisfied.” Codes must be found before they can be evaluated, yet the steps of collecting, evaluating, and deciding may be phenomenologically indistinguishable; that is, the analyst may see a code come up in a string search, recognize almost unconsciously that it is irrelevant, and scroll past it.

As process modelers, one might ask, why should we belabor the point and classify the string search as collection, the barely conscious brush-off as evaluation, and the scrolling past as rejection? If the model were a mere academic conceit, it would indeed be difficult to justify. The justification becomes clear when the model becomes prescriptive rather than descriptive and the steps are broken out in order to perform them in a way that supports the collection of empirical semantics, of metadata that enables reuse. This is covered further in the discussion section. For now, the justification is that these steps (except for reporting) are logically necessary, in the order given, even if they cannot be clearly located in anyone’s lived of observed experience:

- Codes must be found before they can be evaluated;
- Every code found and considered must be either accepted or rejected;
- The complete code set must (implicitly) be judged good enough for use before it can be used; and
- Though reporting does not logically have to happen last, it should — if it happens at all.

#### Contribution 12.

*The work of code set development lies primarily in validation, the iterative collection and evaluation of codes, individually or in groups, and the code set as a whole until it is considered fit for use*.

#### Contribution 13.

*If a reused code set serves only as a starting point, assuming nothing about its quality or its fitness to the current requirements, then the benefits of reuse only affect the code collection process*.

While the evidence we gathered would not support the inclusion of sharing and reuse phases in our process model, we include those activities throughout the code set development process in our recommendations.

### 3.12. Factors that can lead to differences in code sets designed for ostensibly the same clinical phenomenon

Reuse is a harder problem than previously described [9–12,26,27], at least if one accepts that meaningful reuse must provide more benefit than a head start on code collection. The following passage from an investigator and administrator of a large DRN, if accepted, seems to drive a final nail into hopes for meaningful code set reuse:

> Code sets are always context specific. There is no such thing as diabetes in an RWD data source, there might be 50 definitions of diabetes and you have to pick the one that matches your question, data, and methods…We may spend months developing a code set for a specific question, iterating on different algorithms until the investigator is satisfied that the definition matches the needs of the study. (P04-SQ16)

Can there really be a need for 50 different diabetes code sets? If so, could they all be given meaningfully distinct names or descriptions so a potential re-user could effectively choose between them? There are a lot of reasons to expect differences in code sets designed for, ostensibly, the “same” clinical phenomenon. Table 7 categorizes factors that can lead to real or apparent redundancy or inconsistency in code set composition.

**Table 7.** Reasons for code sets to differ

| Valid semantic reasons | Valid empirical reasons | Invalid reasons |
| --- | --- | --- |
| **Clinical meaning or nuance.** Code sets that refer to the “same” clinical idea may actually refer to different ideas sharing an ambiguous name or description.<br>**Study requirements.** Different code sets may share the same clinical meaning, but one study may need a more specific code set, another a more sensitive one.<br>**Algorithmic context.** If a code’s presence causes false positives and its absence causes false negatives, it might be included in some code sets if phenotype or cohort definition rules can be used to eliminate the false positives. Various features of phenotype algorithms can affect the codes needed.<br>**Necessarily arbitrary judgment calls.** If a code’s presence causes false positives and its absence causes false negatives, and this can’t be corrected for in the algorithm, code set authors will be forced to decide whether to include it or not. | **Population characteristics.** E.g., codes may differ for pediatric or geriatric patients; a study of neuropathy in orthopedic surgery patients may exclude diabetic neuropathy.<br>**Regional, institutional, or clinical specialization coding practices.** A single meaning may be expressed using different codes in different places.<br>**Institutional workflow.** Certain conditions or observations may not be captured in some clinical settings requiring recourse to indirect ways of identifying them in EHRs. | **Validation errors.** Codes may be missed in error (potentially causing false negatives) or be included mistakenly (potentially causing false positives.)<br>Reference standards at different sites may be based on identical semantic intentions but differ due to inconsistencies in chart review practices or chart reviewer error. |

Published approaches to discerning the intension^7^ of a code set have been semantically based: identifying a root term or classification and confirming correspondence to that [14,18], and more complex automated vocabulary analysis [19]. Our participants warnings of variations in intension and other factors that can affect code set composition may be exaggerated. Whether the intended meaning and use of a code set — or the differences in intended meaning and use between two code sets — can be parsed out and distinguished from errors and arbitrary decisions remains an open question.

#### Contribution 14.

*A potential code set re-user needs ways to assess both the quality of a candidate code set and its semantic and empirical fitness for their requirements in order for the benefits of reuse to be felt in the processes of code set refinement and validation, acceptance, and validation reporting*.

As an illustration of the assessment needs outlined here, in Appendix B. we provide a list of questions from the point of view of a potential code set re-user.

## 4. Discussion

Code set repositories like the National Library of Medicine’s VSAC provide a helpful service in giving RWD researchers access to code sets they can use as starting points for their own code set development efforts, but greater benefits from reuse have been elusive. VSAC is a vital resource for CQM value sets, but there has not been evidence of uptake in the RWD community. Nevertheless, efforts to facilitate and encourage code set reuse persist year after year. We attribute this persistence to a few aims that we refer to as “the vision of reuse.” Elements of this vision have often been discussed [6,5,12,9,18,19,26,27,23], but seldom if ever operationalized, and the vision persists more as an idyllic intuition than as a program of clear objectives. To briefly unpack the elements of this vision, widespread code set reuse is expected to result in:

- A reduction of duplicative effort,
- An increase in code set quality,
- Progressive refinement of code sets,
- Increasing trust in code set quality; and
- ultimately arriving at standardized code sets that represent common clinical phenomena, so that different studies on different databases will be semantically interoperable at the code set level.

The last of these, standardized code sets, as we have seen, may be impossible or inadvisable as any clinical phenomenon may need multiple code sets and the validation necessary for standardization may need to be performed on phenotype algorithms rather than code sets.

Even if the aim of single, standardized code sets for common clinical phenomena is abandoned, might it be possible to arrive at a small number of widely used code sets for a given clinical idea? If the number of code sets for a given idea could be kept small, it would seem possible to attain the goals of progressive refinement and increasing trust.

If, indeed, 50 different code sets for diabetes could legitimately be needed, and a potential re-user found that many choices in a code set repository, it seems unlikely that they would take the time to sift through them to find the one that matched their use case and the further trouble to refine it and contribute documentation of their use that would increase trust for subsequent re-users. Out of so many candidates, what criteria or methods could be used to match a particular code set to their requirements?

Current tools capture little if any evidence of the validation or evaluation work performed by a code set’s authors or prior users, despite calls for the capture of such evidence [25,26], nor do they tend to capture fine nuances that would distinguish code sets for the “same” concept. Where evidence or results of validation are available, they are not enough to let a potential re-user gauge the applicability of past validation to their own context.

We remain agnostic as to the feasibility of attaining conditions conducive to progressive refinement and increasing trust with RWD code sets. We are, however, optimistic about the potential for innovative tools and further study to bring about the envisioned reduction in duplicative effort and increase in the quality and trust RWD researchers can have in their code sets. This might be achieved without necessarily reusing code sets at all, but by reusing the code-level documentation and metadata that can be captured in the code set authoring process.

### 4.1. Technology and infrastructure recommendations

We agree with CCSE’s [26] emphasis on the importance of code set reuse and the need for the widespread adoption of code set authoring tools that integrate reuse features and that capture the metadata necessary to allow for reuse.

CCSE argues that software for code set construction should, and does not currently, support the validation of code sets. Without saying so explicitly, however, their recommendations seem to suggest that such software should *not* support empirical validation. Particularly Recommendation #2: “Software tools should: 1. be open source, publicly available and easily accessible; […] 3. have minimal setup to facilitate widespread adoption.” [26, p. 8]

We are in strong agreement with these and their other explicit recommendations. Given all the challenges involved in using empirical data and techniques, we are in sympathy with their implied recommendation that software be built that does not rely on these. At the same time, we must recognize that many would not consider a code set validated if it had not been validated empirically.

The tensions between empirical approaches to code set construction and validation and approaches that do not use empirical data are central to the question of whether meaningful reuse of code sets for RWD applications is possible and how.

Hence, we have been at pains to thread two passages through the tensions besetting code set development:

1. In attempting to be meticulous in our conceptual modeling, we hope that software and infrastructure developers will indeed build tools following CCSE’s recommendations, while being mindful of what is gained and what is lost in deciding whether and which empirical techniques will be supported. The OHDSI tools, including ATLAS, assume the presence of empirical data. But could the vocabulary search and concept set functions, which do not rely on PHI, be separated into a detachable module for use by code set developers who may be content using data only in the semantic and empirical semantics categories? All current ATLAS installations are, of course, connected to empirical data sources, and these installations serve OHDSI’s current constituencies. But spinning up a CDM and ATLAS instance is not easy; it can take months. A detachable semantics-only module could be installed and functioning in minutes.
2. In proposing the category of empirical semantics (and, in the paragraphs below, fleshing out how these data can be collected), we envision raising the value of what can be accomplished with semantic-only code set management tools. Perhaps, over time, reservoirs of empirical semantics data will grow, as the tools for collecting and using them mature. We can imagine that even those most committed to MRA-generated reference standard-validated code sets might someday be able to trust some code sets developed without empirical data.

#### 4.1.1. Interfaces for capturing empirical semantics

##### Contribution 15.

*If a code might plausibly be considered for inclusion in a code set, the thought process involved in deciding on it should be captured. To that end, code collection should be separated from code consideration by collecting codes into a separate basket. Each code, singly or in closely related groups, should then be considered and selected for inclusion or rejection, at which point software should prompt for a concise justification of the decision made*.^*8*^

The reasoning behind this recommendation is as follows. More or less every person who has had occasion to think and/or write about code sets has called for better documentation of the code set authors’ intentions. Our general knowledge of human-computer interaction and user interface design tells us that users are annoyed when asked to document their complex thought processes for the benefit of possible future users. Beyond what is conveyed in the name of a code set (e.g., type 2 diabetes milletus, or even type 2 diabetes milletus for screening), the real subtleties involved in crafting a code set arise around factors listed in Table 7. *Reasons for code sets to differ*, and, in our experience, they tend to arise over the course of long conversations involving phenotype definitions, study considerations, and discussants’ knowledge of clinical workflows, population characteristics, and terminology use. None of this is likely to be captured in a metadata field prompted for at the beginning or end of the code set authoring process.

##### Contribution 16.

*By prompting for reasoning at the moment that a single code or group of related codes is actively accepted in or rejected from the code set, the reasoning will be fresh in the authors’ minds, minimizing cognitive load*.

If the reasoning happens to be obvious, the software should facilitate entering it with as little friction as possible, without inadvertently tempting the user to breeze over it when there is something to say. Examples of possible justifications for code rejection:

- Self-evident (e.g., for family history of diabetes);
- Too sensitive (e.g., for gestational diabetes in a code set for a longitudinal diabetes study);
- Frequently used as rule-out code;
- Include only if first of two occurrences in 90 days;^9^
- Drug appears to have been prescribed for a different indication than the one being studied.

By sorting the basket of collected codes into accept and reject categories rather than working on the code set in place, the end point of code consideration becomes clear – the job is done when all codes have been accepted or rejected and the basket is empty. By prompting the user with example justifications like those listed above, software can help them clarify their decision for themselves. It is likely that certain accept/reject justifications will be used more commonly than others, depending on the code set’s purpose and the methods by which codes are collected and considered. Promoting those in the list of prompts may speed the process as the user moves along.

The documentation and metadata captured through this process can be used for validity reporting. These are the metadata mentioned above that can advance the vision of reuse whether or not whole code sets are reused. Metadata on the decisions made to accept or reject specific codes will accumulate and form a body of semantic, sharable evidence, some of it empirically informed, that the current authors and future authors can use and reuse for subsequent code sets.

By structuring the code set authoring process to focus on single or closely related groups of codes, many of the evaluation techniques employed manually and laboriously today could be automated. To be clear, the *evaluation* cannot be automated, what can be automated is the collection and display of relevant information to enrich and optimize the authors’ consideration of the focal code(s). In the ATLAS concept set editor, there is already an ability to click on a single concept and go to a page of information about that code and the codes it is related to, but this can be a cumbersome interface. The user clicks away from their code set to examine the single code. From there further clicks are needed to visit, four separate tabs containing details about the single code, two different views of related codes, and record counts. By the time the user has examined all this and returned to their code set, they may have forgotten what they wanted to know about that code in the first place, or, possibly, even which code they were just inspecting since there is no differentiation on the code set page between codes that have previously been examined and those that have not.

The user interface would ideally allow there to be more than a single focal code, it would expand the scope of information available about the focal code(s), display them without needing navigation across pages and tabs, and conclude the examination with a decision prompt before the user begins to focus on other codes, capturing their decision and reasoning with minimal cognitive burden. If they are not ready to make a decision at that moment, they can be prompted to say what additional information they need before returning the focal code(s) to the holding basket.

The information that should automatically be presented to the user regarding the code(s) under consideration would include:

- Record counts;
- Code sets this code has been used in or rejected from and the justifications given;
- The code(s)’ neighborhood in the semantic graph formed by the source vocabulary;
- The code(s)’ neighborhood in the larger semantic graph formed by the source vocabulary and other vocabularies and ontologies it may be linked to; and
- Machine-generated optimizations and suggestions, such as PHenotype Observed Entity Baseline Endorsements (PHOEBE) [51], which can provide lists of included, excluded, and recommended concepts for code set adjustment.

If the authoring tool did have access to empirical data, it could also show patient records matching the focal code(s) or how the code(s) affected phenotype performance against any available reference standard or registries.

These exploratory tools could reveal semantic subtleties and additional codes that may be worth considering. They could be particularly useful, for instance, in finding existing code sets where the focal code(s), their descendants, or their ancestors had been accepted or rejected.

##### Contribution 17.

*By contributing validation evidence and process metadata to a growing body of empirical semantics, the benefits of reuse might be felt throughout the code set development lifecycle. These benefits may eventually apply to whole code set reuse, as it is conventionally conceived. In the shorter term, they will apply to code-level reuse*.

#### 4.1.2. Expanding the semantic graph

Vocabularies can be structured as semantic graphs. In the case of formal ontologies, these graphs can be used for computational reasoning. These graphs can also be analyzed for insights in other ways, such as interactive visualization.

The OMOP vocabulary system has a simple structure. Dozens of vocabularies and cross-vocabulary mappings are combined into a single concept table and a single concept_relationship table, which together can express most of each vocabulary’s internal semantic structure and the complex semantic structures that arise through linking them. These two tables provide the vertices and edges of a complete semantic graph. All the digital forms of semantic data we have discussed, including empirical semantics, are easily attached to nodes in this semantic graph. We will leave it as an exercise for the reader to imagine the many ways the resulting graph might be navigated, analyzed, visualized, and exploited.

#### 4.1.3. Consolidated recommendations

While we believe the user interface features we recommend above would go a long way toward bringing the vision of meaningful code set reuse into possibility, they do not stand on their own. The software platform we would like to see would combine:

- CCSE’s recommendations [26, pp. 7-9];
- VSAC’s support of standards, versioning, and maintenance following vocabulary updates;
- Recommendations from Gold 2018[9, pp. 6-7];
- ATLAS features, including
  - facet-based search and filtering;
  - integration of code set authoring and sharing with cohort definition and other study algorithms;
  - connection to empirical data for testing of code sets in context; and
  - optimization for parsimonious code set representation;
- PHOEBE’s features for automated code set inclusion/exclusion recommendations; and
- Expert review and approval tracking from N3C’s Concept Set Editor.

### 4.2. Limitations and future work

Our participant sample may not be representative of the field. While intentions were to maximize diversity, we were skewed toward mostly expert users: informaticists and researchers with an interest in RWD study infrastructure in addition to conducting specific RWD studies.

When our participants warned of variations in intention and other factors that can affect code set composition, they did not have specific examples at hand. Further work is needed to explore whether differences in intended meaning and use between two code sets can be parsed out and distinguished from errors and arbitrary decisions.

Questions of code set and phenotype validation need to be better elucidated and more clearly debated in the field, especially in their implications for reuse. We would recommend a special issue of an informatics journal or a dedicated workshop sponsored by a major informatics conference for that purpose. We believe this paper provides a foundation for framing that debate.

## 5. Conclusion

There are two general approaches to validating an analytic code set: (1) semantically, by expert review and approval of the code set’s content, and (2) empirically, by comparing the results of using the code set (in a phenotype or other algorithm) with a reference standard tagged by experts through systematic chart review. There are many reasons to distrust either approach, particularly when reusing a code set in a new study or database context. As long as code set sharing and reuse are separate activities, occurring before and after the main action of code set development, there can be little hope for the vision of reuse. Sharing and reuse must permeate every step of code set development before we begin to see meaningful gains in potential re-users’ trust in the quality and fitness for use of shared code sets and code-level evidence.

## Data Availability

Data produced in the present study are identifiable and private according to the protocol. On reasonable request to the authors, we may be able to produce a deidentified extract.

## 6. Acknowledgments

Richard Williams, Jessica Ancker, Christopher Chute, Davera Gabriel, Harold Solbrig, Jeff Brown, Laura Wiley, Luke Rasmussen, Rachel Richesson, Allen Flynn, Meredith Zozus, David Gotz, Erica Voss, Patrick Ryan, Chrstian Reich, Kristin Kosta, Shelley Rusincovitch, Niklas Elmqvist, Leilani Battle, Joel Chan, Amanda Lazar.

## Appendix A.

### Survey questions

1. Name
2. Email Please include an email address if you might be willing to participate in the next phase of this study, an interview over web conference.
3. Organization, institution, department
4. Title/Role
5. Specializations or degrees related to health care
6. Professional memberships or communities
  ⍰ AMIA
  ⍰ ISPE
  ⍰ OHDSI
  ⍰ HL7
7. If you are a researcher, what are the major topics of your research?
8. What sorts of health data do you work with regularly?
  ⍰ Coded data with codes from standardized terminologies like ICD9, ICD10, SNOMED, MedDRA, NDC, RxNorm, LOINC, etc.
  ⍰ Coded data with codes from local, institutional terminologies
  ⍰ Clinical notes
  ⍰ Radiology images
  ⍰ Clinical trials data
  ⍰ Genomic data
  ⍰ Pathology data
  ⍰ Administrative data
  ⍰ Medical device data
  ⍰ Other (please explain) F*ollow-up:* If your answer does not include coded data, please submit the survey now. (And thank you very much…)
  For the remainder of the survey, please answer the questions specifically as they relate to your work around analyzing or supporting the analysis of coded patient data, that is, data in which clinical events and observations are stored as codes from controlled terminologies.
9. How many substantial studies or analysis projects might you work on in a year?
  ⍰ 0 to 1
  ⍰ 1 to 5
  ⍰ Lots
10. Do you work with teams on these projects?
  ⍰ Yes How many people are on the teams for typical projects? How many organizations participate in your analytic projects? What roles do you play? (E.g., investigator, analyst, statistician, informaticist, clinical expert, epidemiologist, terminologist.) What roles are played by others?
  ⍰ No
11. What data sources and analytic software tools do you and your team use in performing health data analyses?
  a. Data types
    ⍰ Claims
    ⍰ Electronic health records
    ⍰ Clinical trials data
    ⍰ Other. Please list particular products
  b. Data sources
    ⍰ Institutional clinical data warehouse
    ⍰ VA
    ⍰ CMS
    ⍰ Vendor-supplied databases
    ⍰ Synthetic or public datasets like SynPUF or MIMIC
    ⍰ Other. Please list particular products
  c. Data models
    ⍰ OMOP
    ⍰ PCORNet
    ⍰ Sentinel
    ⍰ i2b2
    ⍰ claim forms
    ⍰ local system
    ⍰ Other. Please list particular products
  d. Health data-focused analytic software tools
    ⍰ OHDSI
    ⍰ i2b2
    ⍰ local system
    ⍰ EPIC
    ⍰ Cerner
    ⍰ Oracle Clinical
    ⍰ Other. Please list particular products
  e. General purpose analytic software
    ⍰ R
    ⍰ SAS
    ⍰ SQL database
    ⍰ Tableau
    ⍰ Other. Please list particular products
12. Do code lists play a part in your or your team’s analytic workflow? That is, if you were, for instance, studying statin use in diabetic patients, would your analysis include, in some way, a list of diabetes codes and a list of statin codes? *Follow-up:* If not, please explain and submit survey We will refer to this type of code list as a “clinical concept value set” from here on.
13. Are clinical concept value sets a distinct part of your workflow or an inseparable part of cohort definition and other analytic tasks? *Follow-up:* Please explain
14. Do you create clinical concept value sets, use ones created by others, or both?
15. Are the clinical concept value sets you use limited to a single vocabulary per domain (e.g., SNOMED or ICD10 for diagnoses, LOINC for labs, etc.) or do they draw from multiple vocabularies (e.g., SNOMED and ICD10 for diagnoses)?
16. How do you verify that you have selected the best codes for representing a clinical concept in your analyses?
17. What tools and resources do you make regular use of in developing or working with clinical concept value sets?
18. Do you ever develop clinical concept value sets that you expect or hope will be used by others? *Follow-up:* What do you consider best practices to facilitate discovery and reuse of your value sets by others?
19. What would you change in the tools you use for clinical concept value set discovery, authoring, editing, and evaluation?
20. Do you see a need for new tools for value set and terminology visualization?
21. How would such tools need to fit into your data analytics workflow?

## Appendix B.

### Questions for code set development

- Should I look at other code sets before constructing my own from scratch?
  - Where should I look for existing code sets?
  - What search criteria should I use?
  - What do I do if I find many different code sets that seem to match my criteria?
    - Do I have a way of knowing which ones fit my use case better than others?
    - Can I distinguish one of them to reuse?
    - Should I combine them all into a single list?
  - If I do manage to find and decide on a single or combined code set that matches my criteria:
    - Is it in a data format I can use?
    - Does it use the terminologies I care about?
    - Is it based on versions of the source vocabularies appropriate to my data?
    - Does it use those terminologies consistently with the way my target data use them?
    - Is this code set credible?
    - Is the repository it comes from recognized and accepted in my scholarly/professional community?
    - Am I trying to actually reuse this code set, or do I simply use the codes as a starting point?
      - If starting point, was it worth the work I’ve done to this point or would it have been easier to find these codes in the vocabularies myself?
    - Has any evidence of how the code set was constructed and validated been made available to me?
      - Was consideration and selection of codes influenced in any way by phenotype (or study) characteristics? Or was the code set treated as a self-sufficient algorithm for matching appropriate patient records?
      - If code set performance was checked by review (formal or not) of database results, were those results generated by a containing phenotype algorithm? (Empirical)
      - If code sets are semantically or empirically “validated” separately from the phenotypes they will be contained in, why? Simply because that’s how the process is set up? Because code set authoring and algorithm authoring are different skill sets not usually combined in the same person? Might this be problematic?
      - What database(s) was the code set created for?
      - How did they make inclusion/exclusion decisions?
      - What database(s) was it evaluated with? How?
      - Did they validate against a (gold or silver) reference standard?
        - Do I know how they constructed their reference standard?
          - MRA? How did they ensure accuracy? What training did the abstractors have[36,52]
          - With a random sample of the general database population? A random sample of a subset? How was the subset chosen?
      - Did they use random chart review?
      - What semantic evaluation/validation methods were used?
    - Do I need to do any less validation than I would if building my own code set from scratch?
    - Are there reasons I could trust prior evaluation or validation work, or do I need to do my own from scratch?

## Footnotes

1 By RWD or RCD, we mean data collected in the routine provision of health care and billing. “RWD analysis” is our term of choice for real-world evidence generation, under which we subsume observational research and other analytic uses of RWD.

2 “Phenotype” is an easily misunderstood and sometimes controversial term. In this paper it is used synonymously with cohort definition and analytic variable. All these terms refer to a clinical phenomenon of interest to a researcher (e.g., type 2 diabetes mellitus, exposure to statins) and as shorthand for the algorithms (including vocabulary codes) that identify this phenomenon in a real-word data source. A single RWD will generally require definition and execution of several phenotype algorithms.

3 Code sets are also known as concept sets in the OHDSI [2] and N3C [3,4] communities. They tend to be called value sets in other contexts, such as (common) data element design [5,6] for clinical trials and electronic health records (EHR) systems and clinical quality measures (CQM).

4 Participant IDs are given except in cases where confidentiality might be compromised. When quotes come directly from survey answers, we also indicate the survey question they answered. For the following quotes, the survey question is SQ16 and the participant IDs are P16, P34, P12, and P23. The survey questions text is provided in Appendix A. Survey questions.

5 The distinction here between lexical and semantic is simply between string search on vocabularies, and navigation of their hierarchies and other structures. For simplicity, we consider both to be “semantic”.

6 With the exception of planning. That is, our taxonomy should accommodate any technique involved in directly constructing, validating, and modifying a code set.

7 Intension (with an s) is a term of art. We spell intention with a t when referring to the intentions of humans and intension with an s when referring to a property of a code set, or referring to literature that spells it with an s.

8 CCSE recommends capturing code rejection, and the GetSet application for Read code set authoring does capture code exclusion (which is not the same as rejection.)

9 A phenotype for sleep apnea may require two codes in 30 days, with the first signaling suspected sleep apnea and prompting a sleep study and the second would signaling confirmation; but suspected sleep apnea may be coded as sleep disorder; so a code set for the first instance might include sleep disorder and sleep apnea, while a code set for the second instance might only include sleep apnea.

